# Cost-utility analysis of low-dose pioglitazone in a population with prediabetes and a history of stroke and TIA

**DOI:** 10.1101/2023.11.01.23297949

**Authors:** Fei Yuan, J. David Spence, Jean-Eric Tarride

**Author notes:** Corresponding author: Fei Yuan, MMath Population Health Research Institute, DBCVSRI, 20 Copeland Avenue, Hamilton, ON L8L 2X2, Canada E-mail address.

## Abstract

**BACKGROUND:** Among patients with type 2 diabetes and a history of strokes or transient ischemic attacks, pioglitazone significantly reduces the risk of recurrent stroke. The Insulin Resistance Intervention in Stroke (IRIS) trial found that pioglitazone also reduced the risks of stroke or transient ischemic attacks and new-onset diabetes among patients with insulin resistance. As reported by our previous work, the low-dose pioglitazone was found to provide most of the clinical benefit of high-dose pioglitazone, with fewer adverse effects. We report a model-based economic evaluation to determine the cost-effectiveness of the low-dose pioglitazone versus placebo.

**METHODS:** A lifetime Markov model, with an annual cycle length and five health states (event-free, myocardial infarction, stroke, new-onset diabetes, death), was developed. Transition probabilities were extracted from a subgroup of IRIS patients with insulin resistance, defined by a glycosylated hemoglobin level of 5.7% to 6.4% (mean follow-up of 5 years). Health state costs and utilities were based on public sources and literature data, respectively. Utilities were weighted by time spent in health states to calculate quality-adjusted life years (QALYs). The incremental cost per QALY gained was estimated for the population. Annual discount rates of 0%, 1.5%, and 3% were applied. In addition to deterministic analyses, probabilistic sensitivity analyses were conducted to deal with parameter uncertainty. The analyses were conducted from a Canadian public payer perspective in 2023 Canadian dollars.

**RESULTS:** The base case results indicated that over a lifetime, the expected costs were CAN$31,534 for low-dose pioglitazone and CAN$55,076 for placebo, resulting in a cost saving of CAN$23,542 in favor of the low-dose pioglitazone. Expected QALYs were 25.10 for the low-dose pioglitazone daily and 19.32 for placebo, resulting in a difference of 5.78 QALYs in favor of low-dose pioglitazone. Probability sensitivity analyses with varying discount rates confirmed these results.

**CONCLUSIONS:** Compared with placebo, low-dose pioglitazone is the dominant strategy.

**Key Points:** *Question:* Compared with a placebo, is low-dose pioglitazone cost-effective in treating stroke/ transient ischemic attack (TIA) and new-onset diabetes in a simulated population with prediabetics?

*Findings:* Over a lifetime, the expected costs were CAN$31,534 for low-dose pioglitazone and CAN$55,076 for placebo, resulting in a cost saving of CAN$23,542 in favor of low-dose pioglitazone. Expected QALYs were 25.10 for low-dose pioglitazone daily and 19.32 for placebo, resulting in a difference of 5.78 QALYs in favor of low-dose pioglitazone. Sensitivity and scenario analyses confirmed the results.

*Meaning:* The model-based economic evaluation indicates that low-dose pioglitazone, compared with placebo, is the dominant strategy.

## 1. Introduction

In patients with type 2 diabetes and a history of strokes, pioglitazone was found to significantly reduce the risk of recurrent stroke.^1, 2^ The Insulin Resistance Intervention in Stroke (IRIS) trial, which measured insulin resistance using a homeostasis model insulin resistance score (HOMA-IR), reported that pioglitazone significantly reduced the risk of adverse outcomes such as stroke or transient ischemic attack (Stroke/TIA) and new-onset diabetes on patients with insulin resistance.^3^ This result has been integrated into the American College of Cardiology/American Heart Association (AHA)-2021 guideline for the prevention of stroke in patients with stroke and transient ischemic attack.^4^ As a HOMA score is seldom measured in clinical practice, our previous work therefore studied the effect on a subgroup of patients in the IRIS trial with prediabetes, defined by a glycosylated hemoglobin (HbA1c) of 5.7% to 6.4% or a fasting plasma glucose level of 100mg/dL to 125mg/dL.^5, 6^ Pioglitazone was reported to reduce the risks of myocardial infarctions (MI) and strokes by 43% in 5 years, and reduce new-onset diabetes by 82% over 5 years.^5^ Importantly, the low dose of 15mg daily caused less adverse effects than a high dose (45mg daily) while affording most of the benefit of high-dose pioglitazone.^6^

The long-term health economic implication of pioglitazone, with various doses (15, 30, and 45 mg daily), was studied in a population with diabetes from a Canadian, Britain and German healthcare payer perspectives.^7-9^ All authors found pioglitazone cost-effective compared with either a few first-line therapies or placebo on such a population. However, there are limitations in these investigations. In the study of Coyle et al, treatment effects were measured by life-year-gained instead of qualify-life-year-gained.^7^ This makes it challenging to compare the cost-effectiveness of the drug across different diseases and thus utilize policy-decision marking. None of these authors investigated low-dose pioglitazone in a population with prediabetes and previous strokes or transient ischemic attacks (TIAs). Here, we report the analysis of the cost-utility of low-dose pioglitazone (15mg daily) versus placebo in such a population from a perspective of Canadian public payers.

## 2. METHODS

Using the data collected from the IRIS trial within a median of 4.8 years,^3^ an aggregate Markov model was developed to estimate the costs and effects of treatments in a life-time cost-utility analysis from a perspective of Canadian public payers (Figure 1, Tables S1 and S2). The costs were based on Canadian dollars of 2023; the outcome of effect was measured as quality-adjusted life-years (QALY). A base-case analysis was discounted by 1.5%/year on cost and utility; two scenario analyses were done with either zero or a discount of 3%/year on both items. The analysis and reporting align to the guidelines of Canada’s Drug and Health Technology Agency (CADTH) and the Consolidated Health Economic Evaluation Reporting Standards (CHEERS) reporting guideline.^10^

### 2.1 Interventions

The Insulin Resistance Intervention after Stroke (IRIS) trial was a multicenter and double-blind trial which randomized 3,676 participants with insulin resistance and pre-history of ischemic stroke or TIA from 179 hospitals between 2025 and 2013. It found that pioglitazone with a dose of 15 mg daily being titrated up to 45mg daily, compared with placebo, reduced clinical risks of strokes or myocardial infarctions (hazard ratio, 0.76,95% confidence interval 0.62 to 0.93, p=0.007).^3^ However, adverse effects of the full-dose pioglitazone, such as weight gain, edema and bone fracture, limit the widespread use of pioglitazone. In a study of low-dose of pioglitazone (15 or 30 mg daily) versus placebo based on 1,938 prediabetic participants from the IRIS trial, Spence et al. found that the low-dose pioglitazone was associated with a lower risk of Strokes/MIs (adjusted hazard ratio 0.48, 95% confidence interval 0.3 to 0.76, p = 0.002) and new-onset diabetes (adjusted hazard ratio 0.34, 95% confidence interval 0.15 to 0.81, p = 0.01) compared to placebo. Therefore, the interventions studied here were low-dose pioglitazone versus placebo.

### 2.2 Study population of the health economic evaluation

A hypothetical population with prediabetes and a history of strokes/TIAs consisted of 1,000 participants taking low-dose pioglitazone (8mg or 15mg daily) and another 1,000 taking placebo daily. Sharing as same baseline characteristics as those of the corresponding IRIS subgroup, the mean age of this population is 64 years (Table S3).

### 2.1 Markov model

The aggregated Markov model included five health states: event-free, myocardial infarction (MI), stroke, new-onset diabetes, all-cause mortality (Figure 1). The population of participants entered the model from the health state of “event-free” and exited from the model after they died. Otherwise, they stayed at various heath states. Cost and utility were projected yearly and cumulated by the end of 36 years when the population reached an age of 100 years old. The input parameters and assumptions of the model are in Tables 1 and S2.

#### 3.2.3 Transition probabilities

The clinical treatment effect is expressed as transition probabilities among five health states from either group of treatments. In the IRIS trial, there is a sub-population of 1,520 participants with pre-diabetes and a history of strokes and TIAs, among which 91 on the low dose pioglitazone (6mg, 8mg, 15mg, or 23 mg daily) and 1,429 on placebo. Transition probabilities were extracted from this subpopulation and presented in Table 1.

#### 2.3.2 Measurement and valuation of health

In Table 1, the utility of strokes referred to Allen (2019),^11^ that of myocardial infarctions referred to Lewis (2014), ^12^ and that of new-onset diabetes referred to Zhang (2012).^13^ Utilities were weighted by time spent in health states to calculate overall quality-adjusted life years (QALYs).

#### 2.3.3 Resource use and costs

The unit cost of low-dose pioglitazone referred to that of pioglitazone 15mg extracted from the website of CanadaDrugMart (Table 1).^14, 15^ Resources use and costs were identified for three health states: myocardial infarction, stroke, and new-onset diabetes. Referring to Lamy et al (2022), the Canadian unit costs of related procedures for myocardial infarctions and strokes were originally collected from 2012-05-01 to 2017-07-31 and expressed in the United States dollars.^16^ These unit costs have been converted back to Canadian dollars and inflated to that of year of 2023 using a 2018-01-01 exchange rate and consumer price index (CPI) provided by the Bank of Canada.^17, 18^ The unit cost of newly-occurred diabetes was quoted from the Ontario Medical Advisory Secretariat (2009) and similarly inflated to the dollar of the year of 2023. ^18, 19^ There would not be costs for states of event-free and death.

### 3.2 Analysis and uncertainty

There were three sets of analyses: the reference case analysis with a yearly discount rate of 1.5% on costs and utilities, treated as the main analysis, was compared with scenario analyses for consistency.^10^ Two scenario analyses had a yearly discount with 0% and 3% separately.

Each set of analyses included a deterministic analysis and a probability sensitivity analysis (PSA). The former assumed constant parameters, while the later were used to assess the impact of the uncertainty of the values of parameters on costs and outcomes for treatment. After the Markov model was run 5,000 times, in each of which the model was fed with a set of parameters generated by various random number generators (Table 1), the expected values were obtained by taking the average of mean costs and QALYs from the simulations. Figures S2 and S3 show that the 5,000 simulations were sufficient for incremental mean costs and QALYs to reach a stable point. Transition probabilities were assumed to follow beta distributions, whose two parameters were calculated from transition probabilities taken from the IRIS trial.^5, 6^ Similarly, the utilities of health states, myocardial infarction, stroke and newly-occurred diabetes, were also assumed to follow beta distributions with two parameters calculated from the mean and standard deviation of corresponding states’ utility.^11, 13, 20^ Unit costs of stroke, MI, new-onset diabetes and study medication were assumed to follow gamma distributions.^14, 19, 21^ Parameters of these distributions were calculated based on the means and standard deviation. If their standard deviations of costs were not found, they would be estimated as 25% of the mean value (Table 1).^10^ The analyses reported the costs and QALYs per treatment, increments and incremental cost-effectiveness ratios (ICER).

With the chronic nature of diabetes and secondary prevention of myocardial infarction, stroke, and other cardiovascular diseases, patients were assumed to take the study medication for the rest of their lives. Other assumptions of the model were in Table S2.

Additional scenario analyses were provided with various time horizons and an annual discount of 1.5% on the cost and utility. In Table S5, the Markov model was validated at a 5-year horizon against the data from the IRIS trial.

## 3. RESULTS

### 3.2 Reference case analysis

In Tables 2 and S4, deterministic analyses (DSA) showed that low-dose pioglitazone daily is dominant over placebo. A total summary over 36 years of extrapolation consisted of CAN$55,076 and 19.32 QALYs for the placebo arm, CAN$31,534 and 25.10 QALYs for the low-dose-pioglitazone arm, with a saving of -CAN$23,542 and an incremental QALY of 5.78. These cumulative costs consisted of CAN$46,238 for stroke, CAN$5,260 for MI, and CAN$3,578 for new-onset diabetes spent by the placebo arm, and CAN$26,172 for stroke, CAN$1,682 for MI and CAN$3,673 for new-onset diabetes spent by low-dose pioglitazone daily. The QALYs consisted of 0.36 for stroke, 0.27 for MI, and 3.16 for new-onset diabetes for the placebo, and 0.20 for stroke, 0.09 for MI, and 3.24 for new-onset diabetes in the low-dose-pioglitazone arm. Probability sensitivity analysis (PSA) agreed with DSA that low-dose pioglitazone daily dominated over placebo. A total summary over 36 years of extrapolation consisted of CAN$54,842 and 19.33 QALYs for the placebo arm, CAN$30,237 and 25.30 QALYs for the low-dose-pioglitazone arm, and a saving of -CAN$24,605 and an incremental QALY of 5.97.

The cost-effectiveness plane (Figure 2) showed that most data points scatter in the first and fourth quadrants, with very few in the third quadrant and none in the second quadrant. Cost-effectiveness acceptability curves (Figure 3) showed that low-dose pioglitazone dominated placebo regardless of the choice of values ceiling ratios. The low-dose-pioglitazone arm had a high probability of cost-effectiveness around 85%, even with a willingness to pay (WTP) threshold of CAN$0. It increases to approximately 100% when the WTP threshold is CAN$5000. By contrast, the placebo arm has a low probability of cost-effectiveness around 15% and quickly drops to 0% when WTP threshold increases from CAN$0 to CAN$5000.

### 3.2 Scenario analysis without annual discounts on cost and utility

Deterministic analyses (Tables 2 and S4) have shown that low-dose pioglitazone daily is dominant over placebo. A total summary over 36 years of extrapolation consisted of: a total of CAN$69,746 and 24.31 QALYs for the placebo arm, CAN$41,239 and 32.37 QALYs for the low-dose-pioglitazone arm, and a saving of CAN$28,506 and an incremental QALY of 8.06.

Probabilistic sensitivity analysis (PSA) also showed low-dose pioglitazone dominated over placebo. A total summary over 36 years of extrapolation consisted of CAN$69,650 and 24.32 QALYs for the placebo arm, CAN$40,001 and 32.64 QALYs for the low-dose-pioglitazone arm, and a saving of CAN$29,648 and an increment of 8.33 QALYs.

### 3.2 Scenario analysis with an annual discount of 3% on cost and utility

Deterministic analyses (Tables 2 and S4) showed that low-dose pioglitazone daily is dominant over placebo. A total summary showed a total of CAN$44,600 and 15.74 QALYs for the placebo arm, CAN$24,763 and 19.98 QALYs for the low-dose-pioglitazone arm, and a saving of CAN$19,837 and an incremental QALY of 4.24.

Probabilistic sensitivity analysis (PSA) consistently showed that low-dose pioglitazone dominates over placebo. A total summary over 36 years of extrapolation consisted of CAN$44,578 and 15.75 QALYs for the placebo, CAN$23,924 and 20.15 QALYs for the low-dose-pioglitazone arm, and a saving of CAN$20,654 and an increment of 4.40 QALYs.

### 3.3 Scenario analysis with various time horizons on low-dose pioglitazone and an annual discount of 1.5% on cost and utility

Scenario analyses (Tables 2 and S4) consistently have shown that low-dose pioglitazone daily was dominant over placebo. For a 5-year horizon, a total summary on extrapolation consisted of: a total of CAN$11,787 and 4.43 QALYs for the placebo arm, CAN$4,859 and 4.69 QALYs for the low-dose-pioglitazone arm, and a saving of CAN$6,928 and an increment of 0.27 QALYs.

For a 10-year horizon, there were a total of CAN$21,963 and 8.09 QALYs for the placebo arm, CAN$9,966 and 8.93 QALYs for the low-dose-pioglitazone arm, and a saving of CAN$11,997 and an increment of 0.84 QALYs. For a 20-year horizon, there were a total of CAN$37,977 and 13.67 QALYs for the placebo arm, CAN$18,772 and 16.21 QALYs for the low-dose-pioglitazone arm, and a saving of CAN$19,205 and an increment of 2.54 QALYs.

### 3.4 Validation of the 5-state aggregated Markov model

In Table S5, the life-years and life-year-gained projected by the 5-state aggregated Markov model at a 5-year horizon were compared with those from the IRIS trial. The former projected 4.73 life-year for the placebo arm, 4.98 for the low-dose-pioglitazone one, and an increment of 0.25 life-year-gained. Restricted mean survival time (RMST) was used to estimate the life-year and life-year-gained observed from the IRIS trial: 4.93 life-years for the placebo arm, 5.02 for the low-dose-pioglitazone one, and an increment of 0.09 life-year-gained.^22, 23^ The Markov model estimated a longer life-year-gained (0.16 years) than the IRIS one.

## 4. DISCUSSION

Although pioglitazone is considered as a first-line medication for patients with diabetes, its usage with high doses (such as 45 mg daily) is criticized for various adverse effects. Therefore, it is not yet widely used to prevent stroke and transient ischemic attacks in patients with diabetes and pre-diabetes. Especially, not being included in various public or private insurance plans brings a financial barrier for patients to obtain the medication. As low doses of pioglitazone offer most of the benefit of high doses without causing as much adverse effects, this study specifically focused on the health economic implication of low-dose pioglitazone.

As consistently shown by the reference case and scenario analyses, low-dose pioglitazone daily is found to be dominant over placebo with lower costs and higher QALYs over 36 years of extrapolation. The process of estimation is stable, given that estimated saving in mean cost and incremental QALYs converged well over at 5,000 simulations (Figures S2 and S3). This finding echoes Valentine et al., who reported that 15mg-daily pioglitazone was cost-effective compared with placebo on a UK population with diabetes (an ICER was £5396/QALY-gained). ^8^

The transition probabilities were extracted from a sub-population of IRIS prediabetes participants.^6^ Benefiting from the mechanism of randomization implemented in the IRIS trial, the demographic characteristics of participants distributed roughly balanced from groups of either on low-dose pioglitazone or placebo. This protected the analysis from being distorted by potential confounding factors.

The study has limitations. The effect of low-dose pioglitazone versus placebo was established from features extracted from a subpopulation of the IRIS trial.^6^ The number of participants who took low-dose pioglitazone was small in this population. This leaded to a smaller number of events observed from a health state to another. Consequently, this made it challenging to vividly reflect how patients transferred among health states in the model. Second, the unit cost of low-dose pioglitazone daily was attained from that of generic pioglitazone 15mg from an online pharmacy in Canada.^14^ Depending on retail prices, the yearly unit cost of the medication can vary. Similar issues can appear to the unit costs of outcome events. However, this variation has been covered by the probability sensitivity analyses which showed consistent results across scenarios. Thirdly, subgroup analyses such as on males vs. females would also be worth considering. Moreover, the yearly transition probabilities are fixed as constants over years. They might increase with age. As transition probabilities were extracted from a subpopulation of prediabetes in the IRIS trial, there might not be enough events in subgroups or various age groups for estimating the probabilities. Furthermore, the model was validated by comparing model-projected life-year and life-gained at a 5-year horizon and those from the IRIS trial. For the low-dose-pioglitazone arm, the lifeyears from the two approaches were similar. For the placebo arm, the model-projected life-year was 2.4 months shorter than that from the IRIS trial. This led to a slightly longer life-year-gained (1.92 months) by the model than observed in the IRIS trial. As the IRIS participants were followed slightly less than five years on average, this comparison was approximate but still gave one a sense about the accuracy of the model projection. Costs could not be compared from both approaches, due to the unavailability of multiple occurrences of outcomes from the IRIS trial. However, various scenario analyses showed that the QALY-gained and saving in costs consistently increased with the extended time horizon. Next, the estimation of QALY-gained may be conservative, as the model assumes that participants have the age-independent probabilities of getting various outcome events in each of the 36 years of extrapolation. When the population gets older, it is more likely for the population to experience outcome events. However, our previous work reported that using underestimated transition probabilities in a Markov model can underestimate life-year-gained and QALY-gained.^22, 23^ Thus, the estimation from our model is still conservative. This does not alter our finding on the dominance of low-dose pioglitazone. On the other hand, the dominance of low-dose pioglitazone found here was based on the comparison to placebo in this cost-utility analysis. Coyle et al compared high-dose pioglitazone (30 mg) against metformin (a ICER CAN$54,000/life-year-gained), against glibenclamide (a ICER CAN$2,000/life-year-gained), against diet and exercise (a ICER CAN$27,000/life-year-gained).^7^ Therefore, our next step would be to compare low-dose pioglitazone versus metformin or other alternatives on our target population. Finally, it would be interesting to assess microsimulation models in future research.

## CONCLUSIONS

The results of this model-based economic evaluation based on the IRIS trial indicate that low-dose pioglitazone daily is the dominant strategy when compared to placebo.

## Data Availability

Data is available upon request.

## ABBREVIATIONS

IRIS: Insulin Resistance Intervention in Stroke (IRIS) trial
MI: Myocardial infarction
QALY: Quality-adjusted life-years
TIA: Transient ischemic attack
WTP: Willingness-to-pay threshold
PSA: Probability sensitivity analysis

## SUPPLEMENTARY MATERIALS

The supplementary material for this article can be found online.

## ACKNOWLEDGMENTS

The authors want to acknowledge the investigators of the Insulin Resistance Intervention in Stroke (IRIS) trial for providing access to the data for this investigation.

## DISCLOSURE

The authors declare that they have no conflict of interest.

## FUNDING STATEMENT

This research received no specific grant from any funding agency, commercial or not-for-profit sectors.

---

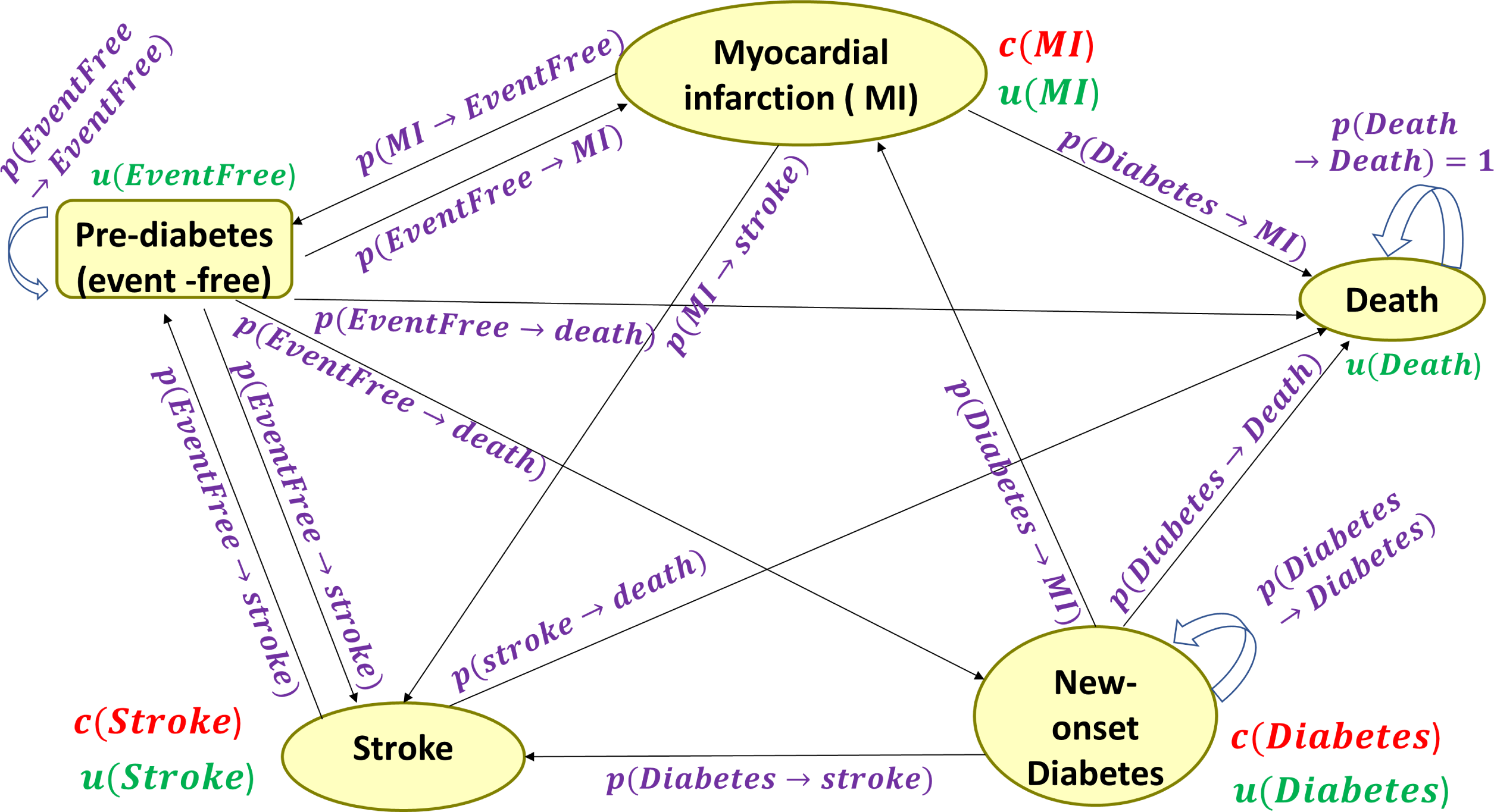

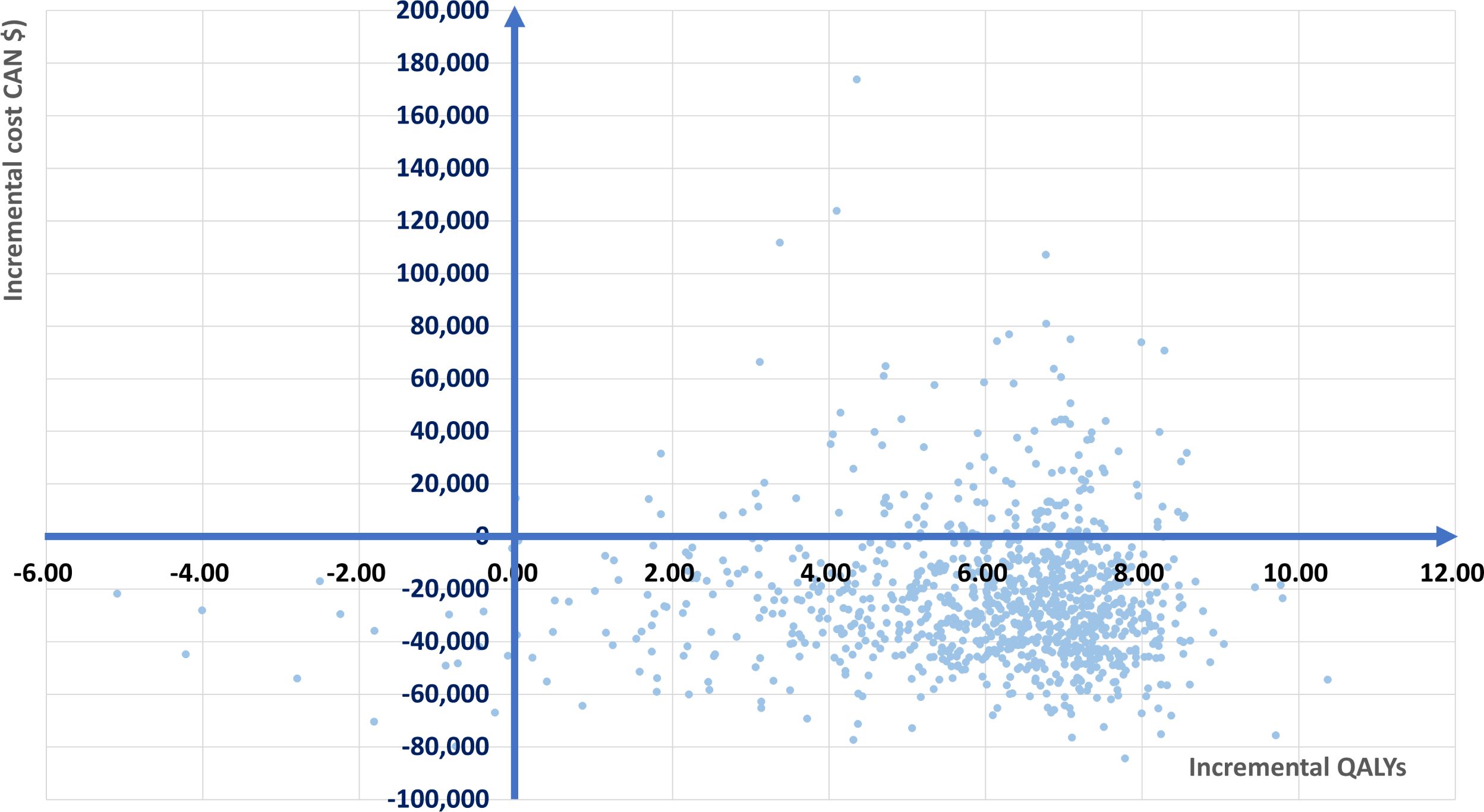

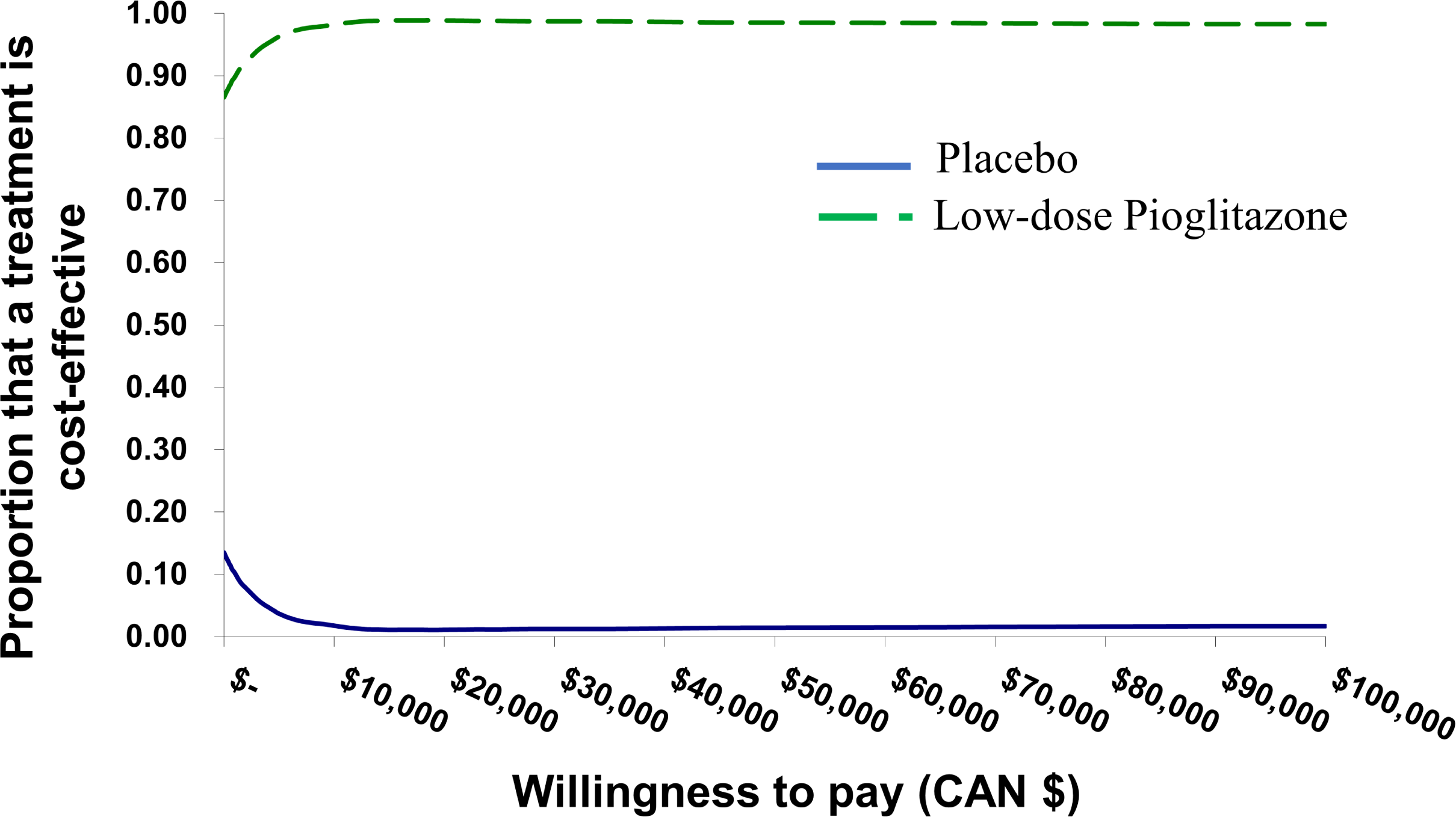

## Notes

### Competing Interest Statement

The authors have declared no competing interest.

### Funding Statement

The manuscript was not supported by any funding resources.

### Author Declarations

The manuscript is not directly related to human subjects. Thus, it does not require IRB approval.

